# High Filtration Efficiency Face Masks made from Sterilization Wraps

**DOI:** 10.1101/2020.05.17.20104638

**Authors:** Sachin Walawalkar, Navin Khattry, B K Sapra, Arshad Khan, Manish Joshi, Lalit Mohan, S P Srivastava, Chital Naresh, Rajendra Badwe, Sudeep Gupta

## Abstract

COVID-19 pandemic has spawned the need for mass production of N-95 respirators. We used Sterilization Wraps to produce face masks which maintained 93% particle capture efficacy post sterilization. This ubiquitously available material could be explored for production of high quality face masks at a cost less than 30 US cents.

## Introduction

Important strategies to prevent SARS-CoV-2 transmission are social distancing and use of personal protective equipment (PPE) such as face masks andN95 respirators [1]. In view of excessive demand for PPEs, hospitals are innovating to protect their staff and patients. Sterilization Wraps, used to wrap sterile surgical instruments, form a formidable barrier to entry of micro-organisms. These wraps are usually made of non-woven polypropylene synthetic fabric consisting of spunbond-meltblown-spunbond (SMS) fibres. These fibres form a matrix that prevents micro-organisms from penetrating the inner layer of the sterilization wrap. Other advantages include the possibility of its reuse after sterilization and fluid resistance. One previous study has reported its use to make face masks [2], which motivated us to explore this material to make face masks and assess their filtration efficiency before and after various sterilization procedures. Our aim was to evaluate the particle capture efficiency of face masks made of this material and the possibility of its use to make N95 respirators.

## Methods

We used sequential sterile wrap, made from Kimguard fabric technology (H300), of2 different metric weights [45 and 60 gram per square metre (GSM)] respectively, to configure a locally made face masks. Each mask comprised 2 layers of GSM 45 or GSM 60, machine stitched to form the outer layer, with the inner layer formed by 2 layers of cellulose hand wipe sheets (figure 1). The inner layer absorbs moisture during breathing to keep the wearer comfortable while the outer layer forms a trap to capture airborne particles in the submicron and micron size ranges. Each mask was 8 cm x 8 cm in its dimensions. Face masks of both 45 and 60 GSM quality were tested for their filtration efficiency pre-sterilization and after sterilization, using 1 cycle of ethylene oxide (EtO) at 37°C, steam at 121°C, heat delivered at a temperature of 69^0^ C with at least 50%humidity for 30 minutes (Thermally Treated), and 2 doses of gamma radiation (5 kGy and 30 kGy), respectively.

**Figure 1:**
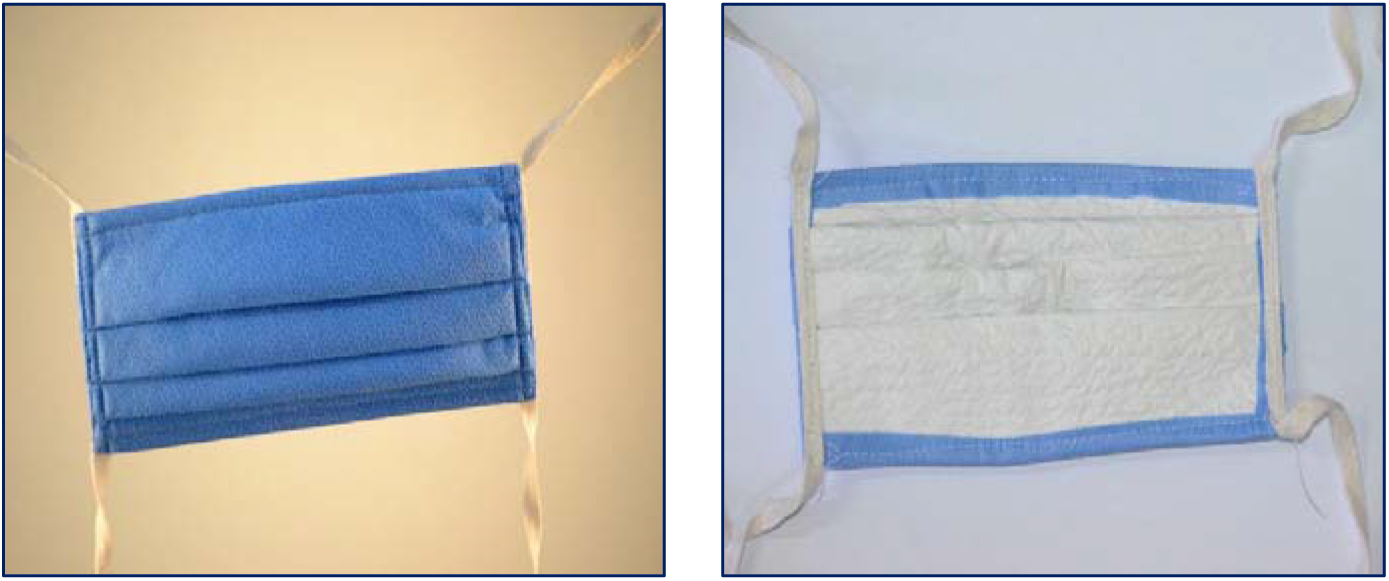
Face mask derived from 60 GSM fabric.

The particle capture efficiency was assessed after cutting the GSM 45 and GSM 60 samples of 1 inch diameter from prototype masks to fit into a filter sample holder meant for aerosol sampling. The test aerosols used were ambient aerosols and the instrument used for the measurements was GRIMM Aerosol Spectrometer 1.108 (Sr No 8F080064) as per ISO 21501-4:2007. The capture efficiency (η) for each size group was estimated as follows:

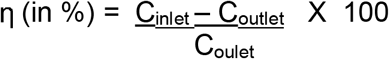

**C_inlet-_** Aerosol number concentration in no/cc at inlet of theaerosol sampler

**C_inlet-_**Aerosol number concentration in no/cc at outlet of the aerosol sampler.

## Results

The results of particle capture efficiency are shown in Table 1. The sample from GSM 60 prototype mask showed a particle capture efficiency of 92.41%at 0.3 micron size in its unsterilized state, which did not reduce after EtO and thermal treatment but diminished after steam sterilization and 30 kGy of radiation. The GSM 45 mask showed a corresponding efficiency of 83.95% in unsterilized state.

**Table 1:** Particle capture efficiency of the filter media for the unsterilized (control) and sterilized face mask samples.

| Particle size (µm) | Aerosol Capture Efficiency for stated size (%) |  |  |  |  |  |  |  |  |  |  |  |
| --- | --- | --- | --- | --- | --- | --- | --- | --- | --- | --- | --- | --- |
|  | GSM 45 |  |  |  |  |  | GSM 60 |  |  |  |  |  |
|  | Control (n=2) | ETO @37 °C (n=2) | Steam @121 °C (n=2) | Thermal treatment at 69°C with humidity (n=3) | Radiation Dose given (kGy) (n=3) |  | Control (n=4) | ETO @37 °C (n=4) | Steam @121 °C (n=4) | Thermal treatment at 69°C with humidity (n=3) | Radiation Dose given (kGy) (n=3) |  |
|  |  |  |  |  | 5 kGy | 30 kGy |  |  |  |  | 5 kGy | 30 kGy |
| **0.3** | 83.95 | 85.97 | 82.00 | 83.63 | 80.55 | 81.09 | 92.41 | 94.48 | 83.98 | 91.47 | 80.44 | 80.05 |
| **0.5** | 92.10 | 94.76 | 91.53 | 95.40 | 93.47 | 94.98 | 97.70 | 98.07 | 92.02 | 98.26 | 92.73 | 94.32 |
| **1** | 96.07 | 96.93 | 98.89 | 100 | 100.00 | 100.00 | 99.45 | 100 | 99.48 | 100 | 100.00 | 100.00 |
| **Total (> 0.3 µm)** | 85.95 | 88.04 | 84.32 | 86.30 | 83.61 | 84.28 | 93.52 | 95.26 | 86.25 | 93.02 | 82.62 | 83.63 |

## Discussion

Our results with a face mask made from a commonly available sterilization wrap material suggest a high filtration efficiency close to the N95 standard. The GSM-60 filter had a higher capture efficiency for 0.3 μm particles as compared to that of GSM 45 filter, both in untreated and after sterilization with ETO and heat at 69^0^ C with humidity, respectively, suggesting the possibility of short-term reuse.

The cost of such face mask in our institution was 30 US cents. Sterilization wrap material made of non-woven polypropylene SMS fibres could be an appropriate readily available inexpensive material for making face masks or N95 respirators.

## Data Availability

All relevant data are within the manuscript and its Supporting Information files.

## FUNDING SOURCE FOR THIS STUDY

None

